# Impact of Stratified Interventions in University Reopenings

**DOI:** 10.1101/2021.08.30.21262805

**Authors:** Yiwei Zhang, Zhuoting Yu, Akane Fujimoto, Pinar Keskinocak, Julie L. Swann

## Abstract

More than 4,000 colleges and universities in the U.S. are scheduled to start a new semester in August or September, 2021. Many colleges require Covid-19 vaccination, as well as some combination of face coverings or diagnostic testing, while others do not (in some cases due to governance structure). Large state universities may especially have limitations and are not requiring vaccination, testing, or indoor face coverings, nor offering hybrid classes (to promote physical distancing). Group living quarters or classrooms with densely packed students are among the riskiest settings for infectious disease spread.

## Introduction

We project cases that may result from the Delta variant of SARS-CoV-2 that is currently circulating widely in the U.S., under various scenarios of initial protection and interventions. Earlier studies of college testing _(1, 2)_ did not consider existing immunity or infections entering the community from external sources, or interventions that may help reduce the transmission (e.g., masks), and previous analysis of K-12 schools _(3)_ did not include different test types or large sizes.

## Methods

We built a Susceptible-Infected-Recovered (SIR) model _(4)_ where vaccinated or previously infected individuals have partial immunity, i.e., move to the Recovered state. The population of 5,000 is well-mixed with 10 additional infections arising weekly from external sources (20 for sensitivity analysis). While some colleges are much larger (e.g., over 30,000 students), it is unlikely that the entire campus simultaneously mixes, so the 5,000 well-mixed population provides a good approximation; the model produces similar results if the initial infections and weekly external infections are scaled proportionally to community size. The model is run for one semester (107 days), and the incoming rate of infections (i.e., the percentage of the population infected at the beginning) is 0.5% (low) or 2.0% (high); if gateway testing is in place then there are no incoming infections. The reproductive rate R_0_ is either 4.0 (baseline) or 5.0 (high), following the genomic surveillance and estimates of increased infectivity of the Alpha and Delta variants of SARS-CoV-2. 40% of the infections are asymptomatic _(5)_, and the average duration of infectivity is 10 days _(6)_. Screening tests have sensitivity and specificity of either (0.7, 0.9) for rapid or (0.85, 0.95) for high-accuracy. A positive test leads to self-isolation, whose length is equivalent to infectivity length. A proportion (33%, 50% or 75%) of all non-isolated individuals are tested weekly regardless of vaccination status. Mask wearing is assumed to reduce infectivity by 50% (universal mask wearing) or 25% (partial mask wearing) _(7)_, where masks are used across all campus settings, including social events. Hybrid classrooms or other distancing reduces R_0_ additionally by 0.5.

## Results

Without masks or surveillance testing in place, in a university with incoming protection of 50%, initial prevalence of 0.5%, the delta variant may infect more than 75% of the susceptible population during the semester. If masks are regularly worn in 25% (50%) of college settings, infections could be reduced to 59% or 24% of the beginning susceptible population. (See Figure 1). Infections rise substantially after about one month. The test positivity on day 30, occurring from screening a portion of the entire population, is indicated on the figure.

**Figure 1:**
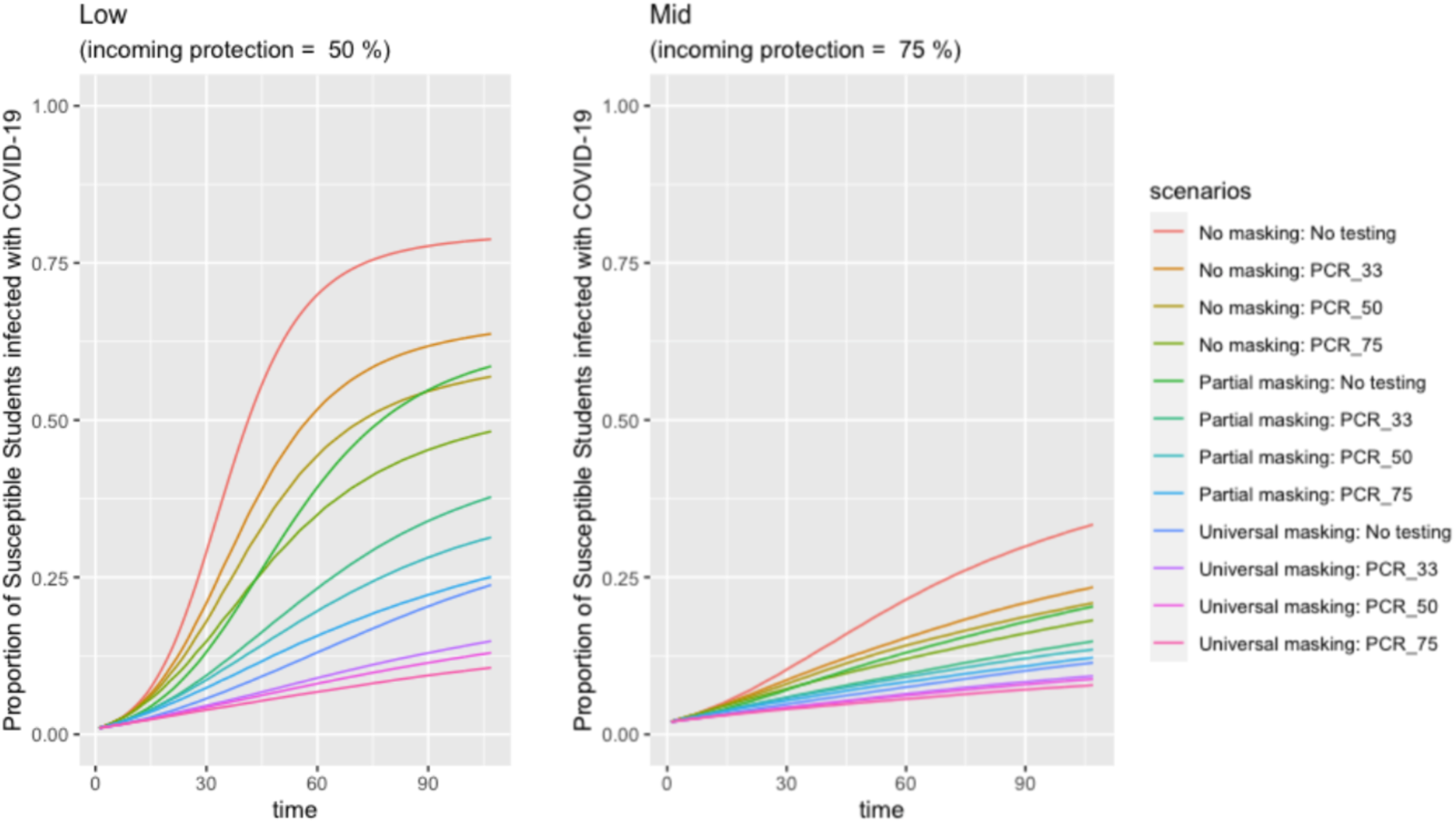
Projected Spread of COVID-19 under different PCR Testing rate and masking strategies. Both of the panels above represent how the proportion of susceptible students infected with COVID-19 increases under certain incoming protection levels (“Low”: 50%; “Mid”: 75%) throughout the semester (107 days). “No masking”, “Partial masking” and “universal masking” scenarios reflect the disease reproduction number of 4, 3 and 2, respectively. Additionally, PCR testing rates of 0%, 33%, 50% and 75% are also included within each setting and masking scenarios.

Screening can be used to get the outbreak more under control. With incoming protection of 50% (75%), weekly testing of 50% of the population with a high-accuracy test results in infections of 57% (21%) of the susceptible population without masks or 13% (9%) of the susceptible population if masks are worn at 50% level.

If Delta is more infectious (R_0_=5.0), 75% of the susceptible population become infected within the semester, even with weekly testing of 75% of the population with a rapid test (or 50% of the population with a high-accuracy test). Only with the addition of masking can the infections be reduced further, down to 19% (See Figure 2).

**Figure 2:**
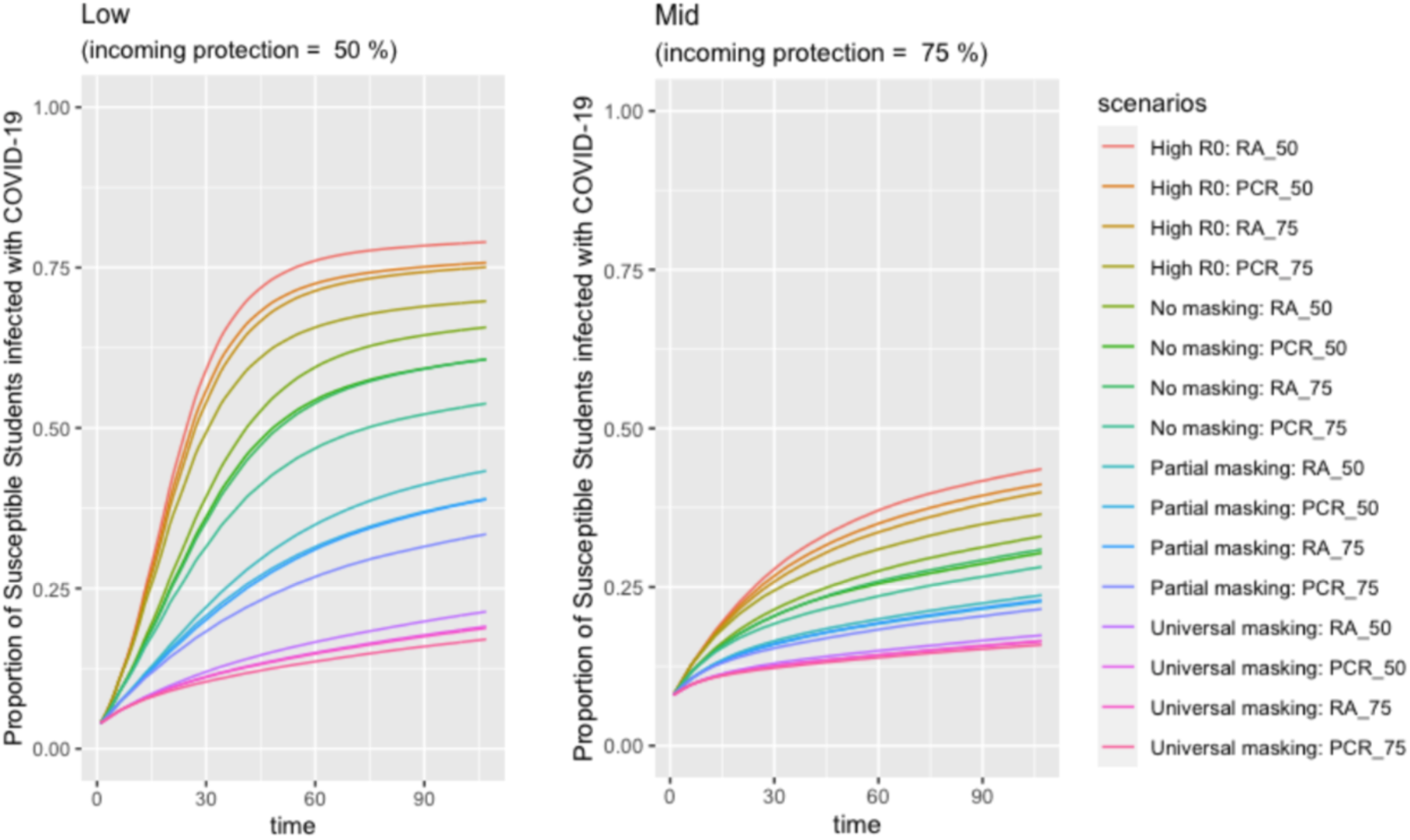
Projection of COVID-19 spread under school testing and masking policies. The panels above represent the proportion of susceptible students infected with COVID-19 with epidemiological settings of two different incoming protection levels. “Low” and “Mid” represent 50% and 75%, respectively. “High R0” reflects the disease reproductive rate of 5. “No masking”, “Partial masking” and “universal masking” scenarios reflect the disease reproduction number of 4, 3 and 2, respectively. Besides, PCR testing and Rapid testing rates of 50% and 75% are also included within each setting and masking scenarios.

If the incoming rate of infections is high (2%), 8% more people get infected compared to the baseline scenario with R_0_=4.0, 25% masks, and 50% PCR testing.

Number of infections is reduced with a higher level of mask usage (e.g., approximately 13% points reduction in infections when mask usage goes up from 25% to 50% in similar scenarios), increased testing (e.g., about 3% fewer infections when the Percentage of population tested weekly goes up from 50% to 75% in similar scenarios), increased testing accuracy (e.g., about 2% fewer infections under high-accuracy versus rapid testing in similar scenarios). Gateway testing also offers benefits (11% fewer infections when gateway testing is in place versus not, in comparable scenarios).

The single best intervention for reducing infections is to have a higher incoming protection level. On average, incoming protection of 75% vs. 50% drops the infections from 48.2% to 27.5% of the susceptible population.

## Discussion

Colleges and universities across the U.S. are facing challenges in keeping Covid-19 cases low in their campus communities. Large public universities have particular challenges as their ability to require masks, vaccination, and regular testing may be limited due to local governance or resource limitations. While encouraging vaccinations can have a significant impact on enabling in-person learning, only 34% of 18-24 year olds were vaccinated by the end of June, 2021 _(8)_.

Several universities do not have accurate knowledge of the vaccination rates or incoming protection (which depends on vaccination, previous disease, and timing) of their community, as their estimates are often based on surveys _(9)_. Universities can compare weekly test positivity to projected values under several scenarios to better understand the current state of a given community, and put interventions in place accordingly to meet desired goals.

Given existing protection levels, mask wearing, and regular testing can reduce infections on campus (and likely surrounding communities). Hybrid classrooms can also reduce infections, though may not be as desirable for the college experience. Masks are likely to be the cheapest intervention though may not be worn in all college settings. Even if a college cannot require it alone, statewide mandates may apply.

Slowing infections on campus will allow additional people to be vaccinated, which largely prevents severe diseases and death. Moreover, slowing spread is imperative in states where hospitals are at risk of being overburdened, which further increases mortality in the community. Employing interventions such as masks, testing, or distancing can help college campuses stay in-person safely, ultimately promoting learning, positive college experiences, and good outcomes in the community.

**Table 1:**
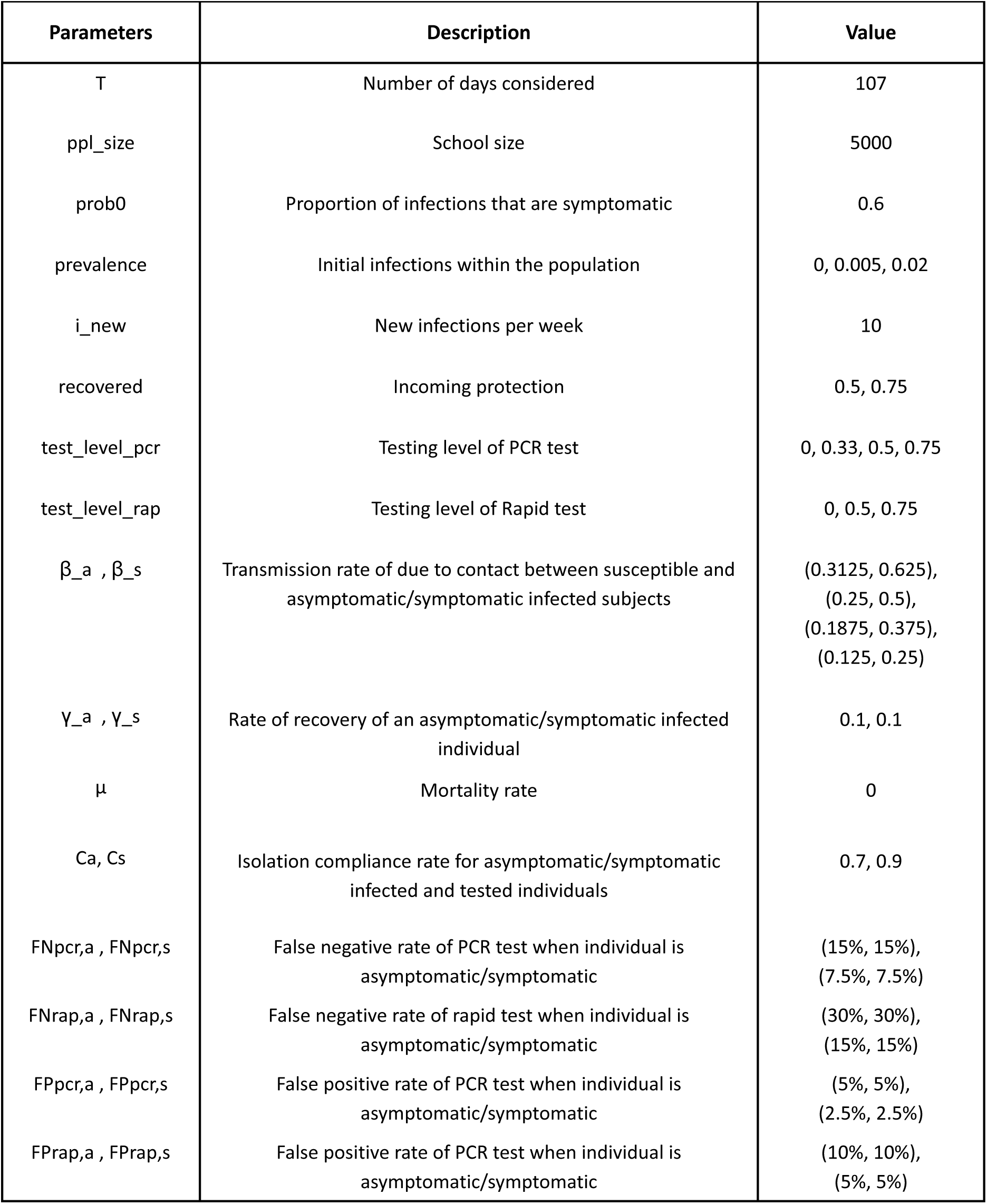
Parameter description and values in the model.

## Data Availability

Data is reproducible with our R code and given parameters.

## References

1. Paltiel, A. D., Zheng, A., & Walensky, R. P. (2020). Assessment of SARS-CoV-2 Screening Strategies to Permit the Safe Reopening of College Campuses in the United States. JAMA Network Open, 3(7), e2016818–e2016818. doi:10.1001/jamanetworkopen.2020.16818

2. Chang, J. T., Crawford, F. W., & Kaplan, E. H. (2021). Repeat SARS-CoV-2 testing models for residential college populations. Health Care Management Science, 24(2), 305–318. doi:10.1007/s10729-020-09526-0

3. Zhang, Y., Johnson, K., Hassmiller Lich, K., Ivy, J., Keskinocak, P., Mayorga, M., & Swann, J. L. (2021). COVID-19 Projections for K12 Schools in Fall 2021: Significant Transmission without Interventions. MedRxiv 2021.08.10.21261726; doi: https://doi.org/10.1101/2021.08.10.21261726

4. Yu, Z., Fujimoto, A. B., Keskinocak, P., & Swann, J. L. (2021). The Impact of COVID-19 Testing on College Campuses. medRxiv,2021.2008.2016.21262153. doi:10.1101/2021.08.16.21262153

5. Oran, D. P. AM, & Topol, E. J. MD. (2020, September 1). ACP Journals. Prevalence of Asymptomatic SARS-CoV-2 Infection. https://doi.org/10.7326/M20-3012

6. Johansson, M. A., PhD, Quandelacy, T. M., PhD, MPH, & Kada, S., PhD. (2021, January 7). SARS-CoV-2 Transmission From People Without COVID-19 Symptoms. Global Health | JAMA Network Open | JAMA Network. https://jamanetwork.com/journals/jamanetworkopen/fullarticle/2774707

7. Mitze, T., Kosfeld, R., Rode, J., & Wälde, K. (2020). Face masks considerably reduce COVID-19 cases in Germany. Proceedings of the National Academy of Sciences, 117(51), 32293–32301. doi:10.1073/pnas.2015954117

8. Baack, B. N., Abad, N., Yankey, D., et al. COVID-19 Vaccination Coverage and Intent Among Adults Aged 18–39 Years — United States, March–May 2021. MMWR Morb Mortal Wkly Rep, 2021;70: 928–933. doi: http://dx.doi.org/10.15585/mmwr.mm7025e2externalicon.

9. Romero, R. (2021). Fake COVID-19 vaccination cards worry college officials. AP News. Available at: https://apnews.com/article/coronavirus-education-fake-vaccination-cards-8c4ca2b4d54434c2fc022b34087df8cb. Accessed on Aug. 18, 2021.

10. Brooks-Pollock, E., Christensen, H., Trickey, A. et al. High COVID-19 transmission potential associated with re-opening universities can be mitigated with layered interventions. Nat Commun 12, 5017 (2021). https://doi.org/10.1038/s41467-021-25169-3

